# A real-world cohort study of immune-related adverse events in patients receiving immune checkpoint inhibitors

**DOI:** 10.1101/2025.01.24.25321082

**Authors:** Yanfei Wang, Yi Guo, Aik Choon Tan, Lili Zhao, Xu Shi, Yong Chen, Ramon C Sun, Mei Liu, Jing Su, Thomas J George, Jiang Bian, Qianqian Song

**Author notes:** **Correspondences:** Qianqian Song, Department of Health Outcomes and Biomedical Informatics, College of Medicine, University of Florida, Gainesville, FL, 32611, Jiang Bian, Department of Health Outcomes and Biomedical Informatics, College of Medicine, University of Florida, Gainesville, FL, 32611.

## Abstract

**Background:** Immune checkpoint inhibitors (ICIs) have significantly improved patient survival outcomes across various cancer types. However, their use is often associated with immune-related adverse events (irAEs), posing challenges in clinical management. Understanding the incidence, severity, and risk factors of irAEs is critical for optimizing ICI therapy and minimizing adverse outcomes.

**Objective:** This study aimed to identify and evaluate risk factors for immune-related adverse events (irAEs) among patients receiving ICIs, focusing on patient demographics, comorbidities, cancer types, and ICI regimens. Additionally, we sought to examine the incidence, severity, and organ-specific patterns of irAEs to guide personalized management strategies.

**Methods:** This retrospective cohort study utilized real-world data from the OneFlorida+ Clinical Research Network, including 9,193 adult patients who received ICIs between January 2018 and December 2022. Patients were categorized based on whether they developed irAEs within one year of initiating ICI therapy. Multivariable logistic regression was employed to identify risk factors for irAEs, adjusting for key covariates such as age, sex, cancer type, smoking status, and comorbidities. Kaplan-Meier survival analysis and cumulative incidence functions were applied to evaluate time to irAE event and overall incidence, stratified by irAE severity, cancer type, and ICI regimens.

**Results:** Of the 6,526 patients included in the final analysis, 56.2% developed irAEs within one year of ICI treatment, including 284 severe cases. Female and younger patients (ages 18-29) were at higher risk of developing irAEs, while comorbidities such as myocardial infarction, congestive heart failure, and renal disease significantly increased irAE risk. In contrast, dementia was associated with a reduced risk of irAEs. Patients treated with combined CTLA4+PD(L)1 inhibitors exhibited a 35% higher risk of irAEs compared to PD-1 inhibitors alone (OR: 1.35, 95% CI: 1.14–1.60, P < 0.001). Cancer type also influenced irAE risk, with breast cancer (OR: 2.36, 95% CI: 1.57–3.60, P < 0.001) and hematological cancer (OR: 2.61, 95% CI: 1.40–5.14, P = 0.004) associated with higher risk compared to melanoma, whereas brain cancer had a reduced risk (OR: 0.55, 95% CI: 0.32–0.92, P = 0.025). Survival analysis revealed that irAE severity significantly impacts both the timing of irAE onset (P = 0.038) and overall survival (P < 0.0001). While treatment regimens significantly influenced irAE-free survival in multi-site cancers (P = 0.02) and overall survival in kidney cancer (P = 0.0011), their effects were less pronounced in other cancer types.

## INTRODUCTION

Immune checkpoint inhibitors (ICIs) have revolutionized cancer treatment, offering unprecedented improvements in patient outcomes across a wide range of malignancies^1^. In particular, ICIs have significantly enhanced survival outcomes, particularly for patients with previously incurable conditions such as advanced-stage non-small cell lung cancer, melanoma, and several other cancers^2-7^. Today, nearly half of all patients with metastatic cancer in high-income countries are treated with ICIs^8^. Currently approved ICIs by the U.S. Food and Drug Administration (FDA) include those that target the programmed death (PD)-1, PD-L1, and cytotoxic T-lymphocyte-associated protein 4 (CTLA-4) pathways, which are crucial in maintaining the immune system’s balance between attacking foreign pathogens and preserving normal tissue. PD-L1, a protein expressed on target cells, binds to PD-1 receptors on CD8 T cells to limit inflammation and protect healthy tissue^9^. However, many tumors express PD-L1 to inhibit T-cell function and evade immune surveillance. ICIs targeting PD-1 or PD-L1 disrupt this immune escape mechanism, allowing cytotoxic CD8 T cells to destroy tumor cells more effectively. CTLA-4, an inhibitory receptor on T cells, downregulates T-cell activity to prevent unnecessary activation^10-12^. Inhibition of CTLA-4 blocks these inhibitory signals, thereby enhancing T-cell activation and immune response against cancer cells^13^.

While ICIs have brought transformative therapeutic benefits, their use has also introduced a new spectrum of side effects known as immune-related adverse events (irAEs), which pose significant challenges in clinical management^14^. Unlike conventional chemotherapy toxicities, irAEs are distinct in their resemblance to autoimmune conditions, yet they follow different natural courses than traditional autoimmune diseases^15^. These adverse events can affect almost any organ system, with the skin, gastrointestinal tract, liver, and endocrine glands being the most commonly involved^16-18^. The severity of irAEs can range considerably, from mild, self-resolving conditions to severe, potentially life-threatening complications^19,20^. The emergence of irAEs necessitates a paradigm shift in clinical practice, requiring oncologists to adopt a more nuanced approach to patient care. Given the complexity and unpredictability of irAEs, clinicians need to monitor these toxicities, ensuring early detection, and implementing timely interventions^21^. Failure to manage irAEs effectively can significantly compromise patients’ quality of life and diminish the overall benefits of ICI therapy^22^.

Despite the increasing use of ICIs across various cancer types, a significant gap remains in the literature regarding the systematic investigation of risk factors that predispose certain patients to develop irAEs, as well as the clinical outcomes associated with these events. Such a knowledge gap limits our ability to effectively predict, diagnose, and manage irAEs. Moreover, a lack of comprehensive studies examining the long-term impact of irAEs on treatment efficacy and patient survival further emphasizes the need for more in-depth research in this area. Several studies have explored different aspects of irAEs associated with ICI therapy, contributing insights while also highlighting the complexity of this emerging field. For instance, Remon et al. focused on irAEs in thoracic malignancies, particularly in non-small cell lung cancer, emphasizing the challenges in managing these events^23^. Valpione et al. identified clinical features and blood biomarkers, such as interleukin-6, as prognostic factors for autoimmune toxicity following anti-CTLA-4 therapy^24^. Eun et al. highlighted risk factors for irAEs associated with pembrolizumab, emphasizing the need for a deeper understanding of these factors^25^. Specific adverse events have also been documented, such as severe neutropenia induced by pembrolizumab, reported by Barbacki et al., which underscores the need to recognize rare but severe irAEs^26^. Delaunay et al. discussed the management of pulmonary toxicity associated with ICIs, underscoring the critical need for specialized approaches^27^. Chitturi et al. evaluated cardiovascular events linked to ICIs in lung cancer patients, highlighting the importance of understanding and managing cardiovascular risks associated with ICI therapy^28^.

While these studies have contributed to our understanding of the diversity of irAEs, they also have notable limitations that need further investigation. Those existing studies are mainly case reports or small cohort studies, which, while insightful, lack the robustness and generalizability to inform clinical practice on a larger scale. On the other hand, systematic reviews and meta-analyses^29-31^ often aggregate data from heterogeneous studies with varying methodologies, leading to conclusions that may lack precision and context. This issue is compounded by the fact that most studies focus on specific cancer types or ICI therapies, limiting their applicability across diverse patient populations. In this study, we aim to address these limitations by conducting a large-scale, systematic investigation of patients who develop irAEs during ICI therapy in a real-world data set. We included different irAE events and cancer types, providing a comprehensive analysis that is both clinically relevant and widely applicable. By identifying key risk factors associated with irAE, our study will contribute to the development of predictive models that can enhance personalized medicine approaches in oncology.

## METHODS

### Study Design

This retrospective cohort study used real-world data from the OneFlorida+ Clinical Research Network^32^, covering ~17.2 million patients from Miami, Orlando, Tampa, Jacksonville, Tallahassee, Gainesville, and rural areas of Florida^33^. The OneFlorida+ data encompasses a broad spectrum of patient data, including diagnoses, procedures, observations (e.g., vital signs), prescriptions, and laboratory results. The study focused on adult patients (≥18 years old) who received at least one cycle of FDA-approved ICI therapy, including PD-1 inhibitors (Nivolumab, Pembrolizumab, and Cemiplimab), PD-L1 inhibitors (Atezolizumab, Durvalumab, and Avelumab), and the CTLA-4 inhibitor Ipilimumab. The study period spanned five years, from January 1, 2018, to December 31, 2022. To ensure data integrity, patients were excluded if they had missing treatment dates or if death occurred during the treatment period, preventing the completion of at least one ICI cycle. This study has been approved by the University of Florida Institutional Review Board (IRB# 201702876).

### Indicator Variables

We included data on age at the time of the first ICI treatment, sex, race and ethnicity, smoking status, and body mass index (BMI). BMI categories were defined according to standard thresholds. Smoking status was defined based on patients’ history. Comorbidities were identified using International Classification of Diseases, Ninth/Tenth Revision, Clinical Modification (ICD-9/10-CM) codes. These comorbidities were recorded as binary variables, indicating the presence or absence of the condition prior to the initiation of ICI. This approach allowed us to standardize the assessment of comorbid conditions across the cohort, ensuring consistency in evaluating the potential impact of pre-existing health conditions on irAE. Primary cancer types were identified using our prior cancer diagnosis algorithms^34-36^. In cases where a patient had multiple cancer types, the diagnosis was categorized under multi-site malignancies^34-36^.

### Outcomes

The primary outcome of interest was the occurrence of irAEs following the initiation of ICI therapy. Identification of irAEs was conducted using a comprehensive list of adverse event codes, as depicted in **Supplementary Figure 1**, where the corresponding irAE codes can be found^37^. Patients were categorized into two groups: the irAE group, consisting of those who developed irAEs within one year of initiating ICI therapy, and the non-irAE group, which included patients who did not experience irAEs. To minimize the risk of underreporting due to insufficient follow-up, the non-irAE group was further limited to individuals with a documented follow-up visit at least one year after their initial treatment. Severe irAEs were defined as cases where patients experienced irAEs and required hospitalization within one month of ICI therapy.

### Statistical Analysis

Descriptive statistics summarized the baseline characteristics of the study population, with counts and percentages presented. Fisher exact test was utilized to compare categorical variables between patients with and without irAE, with p-values calculated to identify statistically significant differences. Multivariable logistic regression models were constructed to evaluate the risk factors associated with irAE. These models adjusted for key covariates, including age group at ICI initiation, sex, race and ethnicity, smoking status, primary cancer type, type of ICI treatment, and comorbidities. Adjusted odds ratios (ORs) with 95% confidence intervals (CIs) were reported to quantify the association between these covariates and irAE risk.

Kaplan-Meier analysis was performed to estimate the probabilities of time-to-irAE, which was defined as the duration from the initiation of ICI therapy to the first irAE event, death, or the last follow-up, whichever occurred first. Patients who did not experience an irAE were censored at the time of death or their last follow-up. Log-rank test evaluated the differences of time-to-irAE between subgroups of different cancer types and different irAE severity (non-severe vs. severe). Cumulative incidence functions were also utilized to analyze irAE incidence. Fine-Gray method was applied to estimate the cumulative probability of developing irAEs over time, while Gray’s test compared cumulative incidence curves between subgroups.

## RESULTS

### Cohort Collection

The study cohort was drawn from the OneFlorida+, comprising 16,936 patients undergoing ICI treatment. 7,743 patients were excluded due to the presence of pre-existing immune-related conditions, resulting in a final eligible cohort of 9,193 patients. Within this cohort, 3,665 patients developed irAEs within one year of ICI treatment, while the remaining 5,528 were classified as non-irAE patients. To ensure adequate follow-up, the non-irAE group was further restricted to 5,528 without irAE within one year of ICI individuals with a documented follow-up visit at least one year after ICI treatment, resulting in a refined non-irAE group of 2,861 patients. Consequently, the final analysis included a total of 6,526 patients, with an overall irAE incidence rate of 56.2% in the study population. The selection and exclusion process for the final cohort is detailed in **Figure 1**.

**Figure 1:**
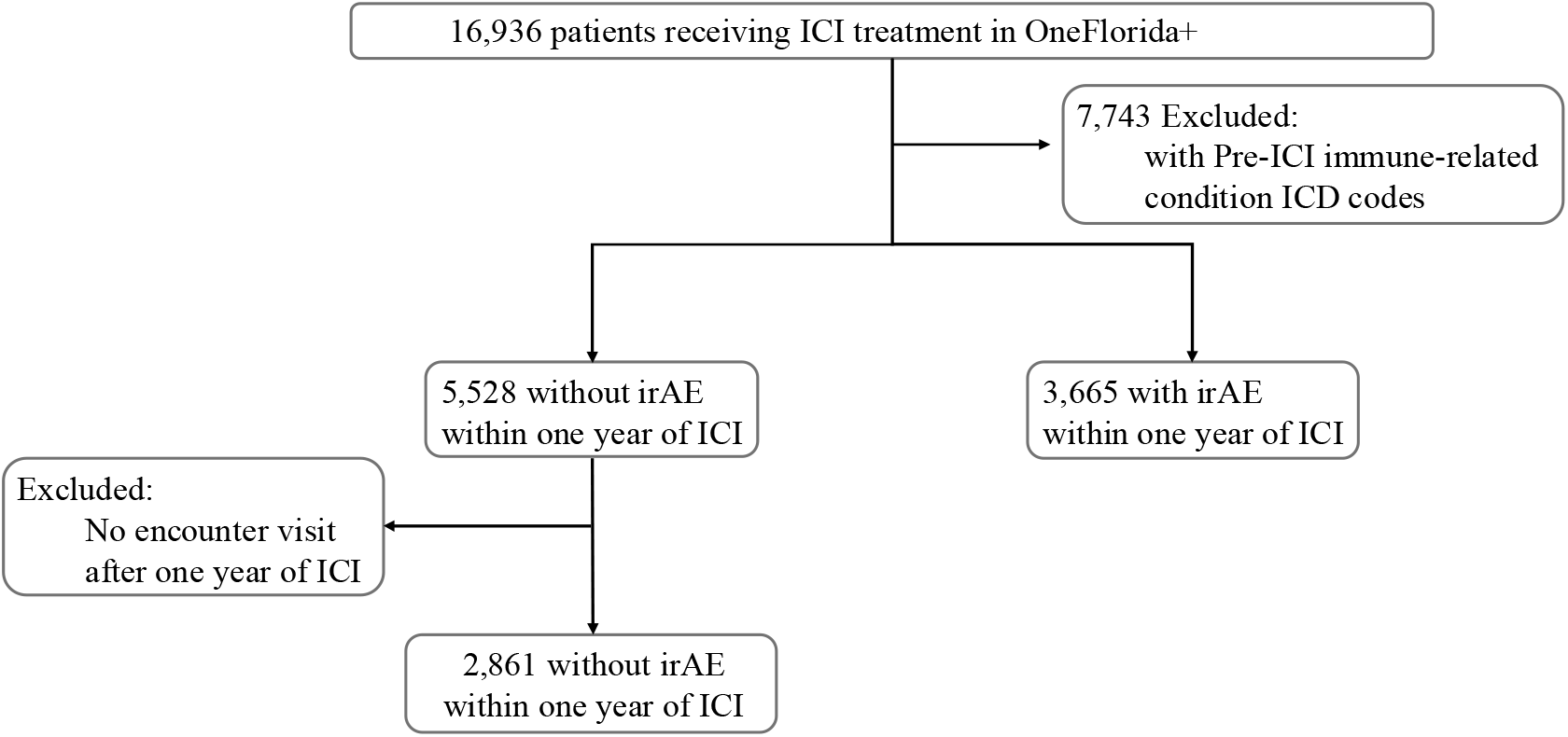
Study Participant Flowchart for ICI Cohort.

### Basic Characteristics

In this study cohort, we collected data on 6,526 patients who received immune checkpoint inhibitors (ICIs), of whom 3,665 experienced irAEs, and 284 had severe irAEs (**Table 1**). Among the patients who developed irAEs, 58.3% were male, compared to 61.7% in the non-irAE group, indicating a slightly higher incidence of irAEs among females. The age distribution of the entire cohort was skewed towards older adults, with 88.3% of patients being over 50 years old. Regarding race, most patients were White (84.8%), followed by African Americans (12.4%). Hispanic ethnicity was less prevalent, with 5.6% of the irAE group identifying as Hispanic, compared to 7.1% in the non-irAE group. Individuals with a history of smoking had a higher incidence of irAEs (7.0%) and severe irAEs (6.3%) compared to those who never smoked (‘Never’), who had a lower incidence of irAEs (5.9%) and severe irAEs (2.1%), suggesting a potential link between smoking and the development of irAEs. 4.3% of irAE patients were classified as underweight (BM/ < 18.5), compared to 3.6% in the non-irAE group. The distribution of patients across other BMI categories—normal(18.5 BM/ < 25), overweight(25 BM/ < 30), obesity(30 BM/ < 40), morbid obesity (40 BM/) —was comparable between the irAE and non-irAE groups.

In terms of comorbidities, myocardial infarction was observed in 7.7% of irAE patients compared to 3.5% in the non-irAE group, with a significant difference (*P* < 0.001). Congestive heart failure also showed a significant difference, with 9.4% in the irAE group versus 4.6% in the non-irAE group (*P* < 0.001). Liver disease was prevalent in 29.2% of irAE patients and 27.1% of non-irAE patients, with a *P* value of 0.058, indicating a trend towards significance. Renal disease showed a significant difference, with 16.3% in the irAE group versus 12.0% in the non-irAE group (*P* < 0.001). Further analysis of cancer types revealed that, aside from multi-site cancers, melanoma was the most prevalent, occurring in 26.0% of patients who experienced irAEs, compared to 20.3% in those who did not. Among those 744 irAE melanoma patients, 557 patients received anti-PD-1 therapies while 149 were treated with anti-CTLA4-anti-PD(L)1. Less common cancer types have been grouped under ‘Other Cancers,’ with additional details provided in **Supplementary Table 1**.

We further explored the 3,665 irAE patients based on the specific organs affected, as shown in **Supplementary Table 2**. Multi-site irAEs were the most common, affecting 1,638 patients (44.7%), followed by gastrointestinal irAEs in 927 patients (25.3%) and neurologic irAEs in 258 patients (7.0%). Hematologic irAEs were observed in 192 patients (5.2%), while renal and respiratory irAEs affected 172 (4.7%) and 167 (4.6%) patients, respectively. Interestingly, over 70% of patients with cardiac, renal, and respiratory irAEs were male. Comorbidities played a significant role in the development of organ-specific irAEs. For instance, 25.3% of patients with cardiac irAEs had a history of myocardial infarction, and 25.3% also had liver disease, while 30.3% of these patients had diabetes. Patients with a history of liver disease accounted for 29.2% of those with hematologic irAEs and 42.2% of those with hepatic irAEs. Additionally, 42.3% of patients with reproductive irAEs had liver disease, highlighting its significant contribution to specific irAE types. Furthermore, 27.3% of patients with renal irAEs had diabetes, and chronic pulmonary disease was present in 45.5% of those with respiratory irAEs, suggesting that pre-existing conditions may predispose patients to organ-specific irAEs.

Cancer type and the type of ICI therapy played crucial roles in determining the organ involvement in irAEs. In melanoma patients, endocrine irAEs were notably more common among those treated with PD-1 inhibitors (33.6%) compared to those receiving PD-L1 therapies (0.84%), with a statistically significant difference (p = 0.00739). Similarly, kidney cancer patients treated with PD-1 inhibitors exhibited a significantly higher incidence of multi-site irAEs (43.0%) than those treated with PD-L1 therapies (3.56%, p=2.55e-26). Conversely, hematologic irAEs were more frequent in patients treated with anti-PD-L1 therapies, particularly among those with non-melanoma skin cancer, suggesting potential therapy-specific risks for blood-related irAEs in this population. These findings underscore the importance of considering both cancer type and ICI therapy when managing irAE risk, as different checkpoint inhibitors may provoke distinct immune responses that impact organ systems variably across cancer types.

### Risk factors for immune-related adverse events in ICI therapy

We first investigated associations between patient demographics and the development of irAEs within one year of receiving ICIs. As illustrated in **Supplementary Figure 2**, female patients exhibited higher risks of developing irAEs compared to male patients (OR: 1.17, 95% CI: 1.06-1.30, *P-value* = 0.002). Age was also a significant factor; younger patients (18-29 years) demonstrated a markedly higher risk (OR: 2.04, 95% CI: 1.26-3.40, *P-value* = 0.005) relative to those aged 65 and above. Hispanic ethnicity was associated with a decreased risk of irAEs (OR: 0.88, 95% CI: 0.79-1.00, *P-value*=0.043). Additionally, smoking status affected the incidence of irAEs, with ever smokers exhibiting a higher risk than never smokers (OR: 1.90, 95% CI: 1.42-2.54, *P-value* < 0.001). Patients classified as underweight (BM/ < 18.5) had the highest risk of developing irAEs compared to the normal BMI group (18.5 BM/ < 25), with an OR of 1.25 (95% CI: 0.96–1.64; *P-value* = 0.099). This was followed by patients with obesity (30 BM/ < 40), who had an OR of 1.05 (95% CI: 0.90–1.22; *P-value* = 0.537), and those who were overweight (25 BM/ < 30), with an OR of 1.02 (95% CI: 0.91–1.16; *P-value* = 0.696). In contrast, morbid obesity (40 BM/) was associated with a reduced risk of irAEs (OR: 0.83; 95% CI: 0.69–1.01; *P-value* = 0.061).

We incorporated comorbidity data into the logistic regression analysis, to further explore health conditions on the risk of developing irAEs (**Supplementary Figure 3**). Notably, patients with a history of myocardial infarction had a 73% increased risk of irAEs (OR: 1.73, 95% CI: 1.35-2.23, *P-value* < 0.001), while those with congestive heart failure showed a similarly elevated risk (OR: 1.74, 95% CI: 1.40-2.17, *P-value* < 0.001). Renal disease also emerged as a significant predictor, associated with a 35% higher risk of irAEs (OR: 1.35, 95% CI: 1.16-1.57, *P-value* < 0.001). Conversely, certain conditions were linked to a reduced risk of irAEs. For example, patients with dementia were found to have a 45% lower risk of developing irAEs (OR: 0.55, 95% CI: 0.36-0.84, *P-value* = 0.006). Additionally, the type of ICI treatment administered played a critical role in the development of irAEs (**Supplementary Figure 4**). Patients treated with a combination of CTLA4 and PD-1 inhibitors exhibited a 35% higher risk of irAEs (OR: 1.35, 95% CI: 1.14-1.60, *P-value* < 0.001), compared to those treated with PD-1 inhibitors alone. Similarly, patients receiving PD-L1 inhibitors also showed an increased risk, with a 28% higher risk of irAEs (OR: 1.28, 95% CI: 1.09-1.50, *P-value* = 0.002).

As illustrated in **Figure 2**, cancer type significantly modulated the risk of developing irAEs among patients undergoing ICI therapy. Compared to melanoma, which served as the reference group, several cancer types were associated with a substantially higher risk of irAEs. Patients with breast cancer had more than double the risk (OR: 2.36, 95% CI: 1.57-3.60, *P-value* < 0.001), while those with secondary malignant neoplasms exhibited an even higher risk, with nearly a threefold increase (OR: 2.92, 95% CI: 1.69-5.27, *P-value* < 0.001). Similarly, hematological cancer was associated with a more than two-and-a-half-fold increase in risk (OR: 2.61, 95% CI: 1.40-5.14, *P-value* = 0.004). In contrast, patients with brain cancers demonstrated a significantly reduced risk of developing irAEs (OR: 0.55, 95% CI: 0.32-0.92, *P-value* = 0.025). This reduced risk may be explained by the high mortality rate in this population, potentially limiting the time for irAEs to manifest. However, other factors, such as differences in immune response or treatment protocols, may also contribute to this observation. To eliminate the potential impact of gender on specific irAEs, we excluded all patients who experienced reproductive irAEs (all of whom were female). We found that the results remained consistent with those obtained without this exclusion, as shown in **Supplementary Figure 5**.

**Figure 2:**
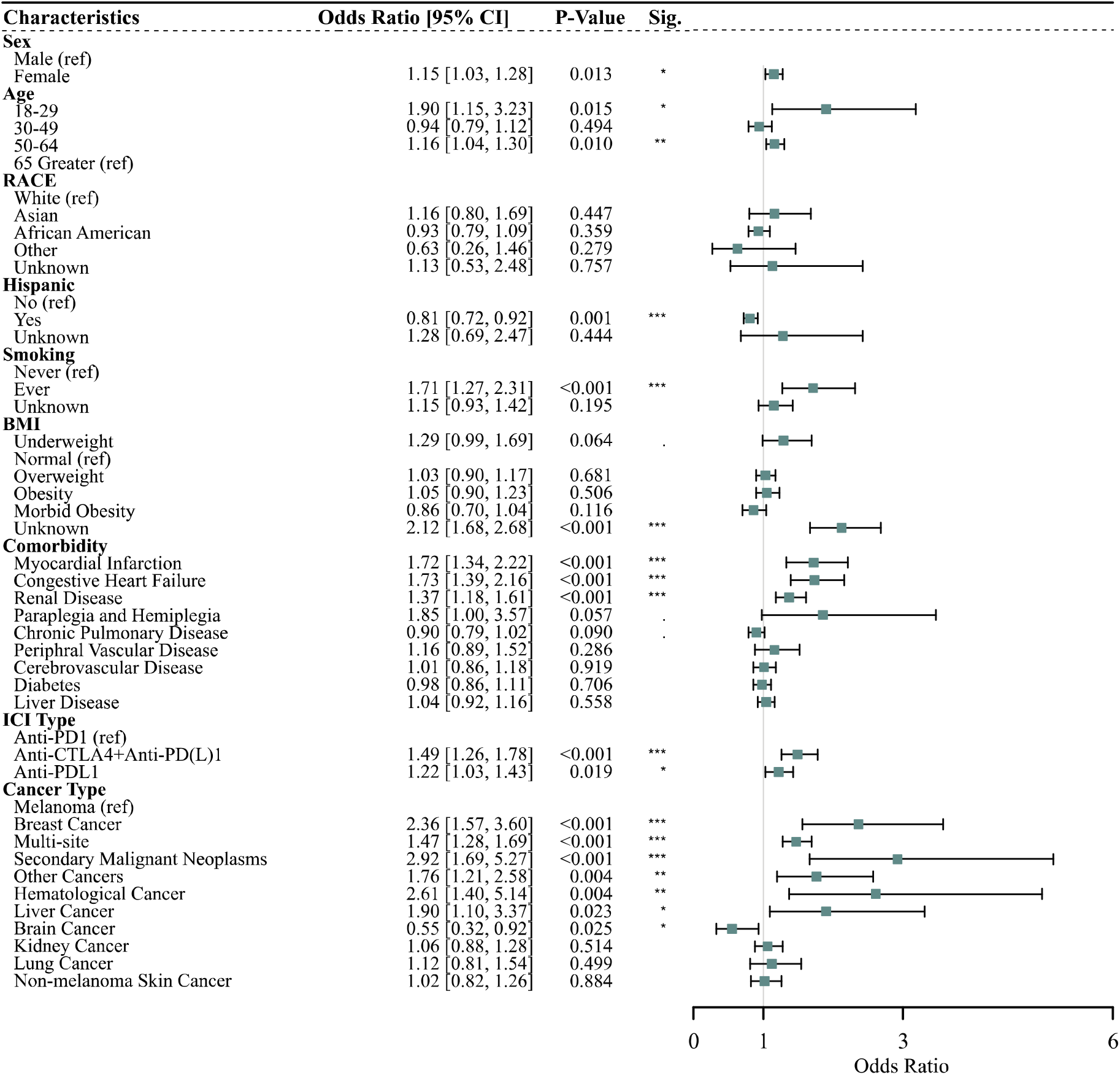
Multivariable logistic regression forest plot for irAE.

### Survival Analysis for irAE free and overall survival

We examined the time to irAE event and overall survival probabilities across different patient groups, stratified by irAE severity and treatment regimens (Anti-CTLA-4 + Anti-PD-(L)1, Anti-PD-1, and Anti-PD-L1). **Figure 3A** illustrated irAE-free survival, revealing a statistically significant difference (P = 0.038) in the time to irAE onset between patients with severe and non-severe irAEs. Patients with severe irAEs experienced a shorter irAE-free survival, indicating an earlier onset of adverse events during ICI therapy. Correspondingly, **Figure 3C** demonstrates a highly significant difference in overall survival between severe and non-severe irAE groups (P < 0.0001). These results collectively highlight that irAE severity not only influences the timing of adverse events but also profoundly impacts long-term survival outcomes, underscoring the clinical importance of effectively managing severe irAEs to optimize therapeutic benefits.

**Figure 3:**
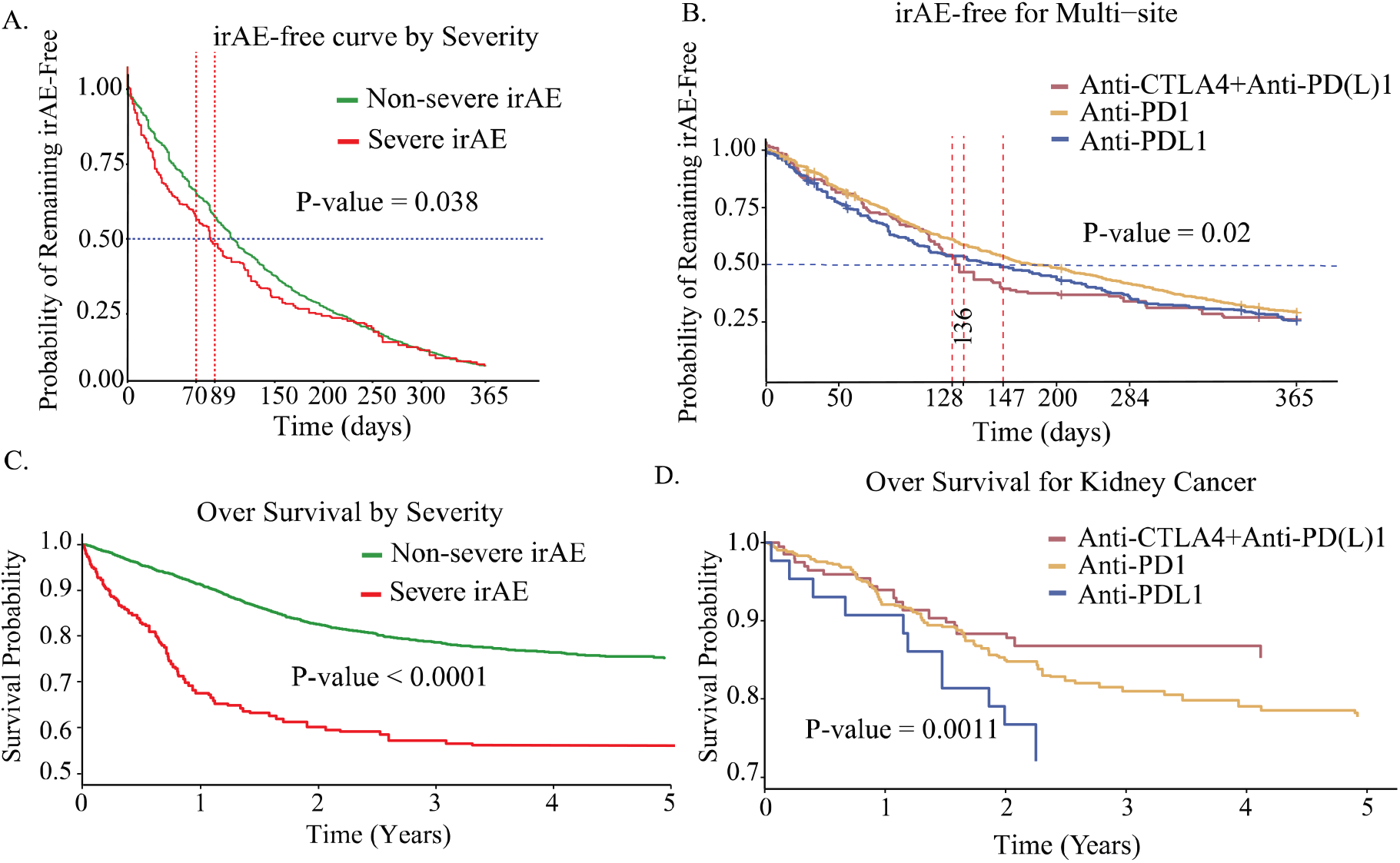
Kaplan-Meier Analysis.

Among these groups, multi-site cancer patients exhibited a significant difference in irAE-free survival across treatment regimens (**Figure 3B**, P = 0.02), suggesting that treatment choice plays a key role in determining the timing of irAE onset in this population. In contrast, irAE-free survival differences were not statistically significant for kidney cancer patients (P = 0.14; **Supplementary Figure 6B**) or melanoma patients (P = 0.1; **Supplementary Figure 6A**), implying a limited influence of treatment regimens on irAE timing in these groups.

Regarding overall survival, no statistically significant differences were observed across treatment regimens for melanoma patients (P = 0.6; **Supplementary Figure 6C**) or multi-site cancer patients (P = 0.83; **Supplementary Figure 6D**). These findings suggest that while treatment regimens may affect irAE-free survival in multi-site cancers, their influence on long-term survival outcomes remains limited. In contrast, kidney cancer patients exhibited a highly significant difference in OS across treatment regimens (P = 0.0011; **Figure 3D**), emphasizing the substantial impact of treatment selection on long-term survival outcomes in this subgroup.

To further understand the dynamics of irAE onset, we examined the cumulative incidence of irAEs stratified by cancer type and severity. **Supplementary Figure 7A** shows that patients with severe irAEs had significantly higher cumulative incidence rates over time compared to those with mild irAEs (P = 0.0175). This supports the observation that severe irAEs occur earlier and at a higher rate during ICI therapy, underscoring the importance of proactive monitoring and early intervention for high-risk patients. Additionally, cumulative incidence curves stratified by cancer type (**Supplementary Figure 7B**) provide insights into the role of cancer burden in irAE development. Patients with multi-site cancers demonstrated a notably higher cumulative incidence of irAEs compared to melanoma patients, with the difference being highly significant (P = 0.0000546). This suggests that patients with multi-site cancers are more susceptible to irAEs, potentially due to the broader immune activation associated with multiple tumor sites. The steeper rise in the cumulative incidence curve for multi-site cancer patients indicates that irAEs develop more rapidly and at a higher frequency in this group. These findings emphasize the need for closer clinical surveillance and tailored management strategies for patients with extensive cancer involvement to mitigate the risks of irAEs.

## DISCUSSION

This study provided a comprehensive analysis of irAE risk factors in patients receiving ICI therapy. Our analyses encompassed patient demographics, comorbidities, cancer types, and ICI treatment regimens. First, we identified that female patients were at higher risk for irAEs, possibly due to hormonal differences and genetic factors that enhance immune activation^38,39^. Similarly, younger patients also exhibited higher irAE rates, suggesting that immune anergy, or the reduction in immune reactivity, may develop with age, thereby lowering irAE susceptibility in older individuals. Our study further observed that certain comorbidities, such as cardiovascular conditions, significantly influence irAE risk. For instance, patients with a history of myocardial infarction or congestive heart failure were more likely to develop irAEs.

This finding aligned with earlier research suggesting that pre-existing cardiovascular conditions can exacerbate immune responses during ICI treatment, potentially increasing inflammation and irAE risks^40-42^. In contrast, an intriguing finding from our study was the reduced risk of irAEs in patients with dementia. While the underlying mechanisms remained unclear, this observation may suggest chronic immunosuppression or altered immune signaling in dementia patients, reducing susceptibility to irAEs. Alternatively, it is possible that irAEs are underreported or masked in dementia patients due to overlapping symptoms with the disease itself or diagnostic challenges. Further research is needed to unravel the complex interplay between neuroinflammation, immune checkpoint inhibition, and irAE risk in this population.

Our findings also revealed that the type of ICI therapy significantly influences irAE risk. Patients receiving combined CTLA4+PD(L)-1 inhibitors showed a higher incidence of irAEs compared to those treated with PD-1 or PD-L1 inhibitors alone. This finding aligned with existing literature^15,43^, which has consistently reported increased toxicity with combination therapies due to the synergistic effects of dual immune checkpoint blockade^44^. Additionally, cancer type significantly modulated the risk and type of irAEs. Specifically, patients with hematological cancers and breast cancers exhibited a substantially higher risk of irAEs compared to those with melanoma. Prior studies have suggested similar trends, particularly for hematological malignancies, where the immune system is inherently more involved, making these patients more susceptible to irAEs^45,46^. Furthermore, our study uncovered distinct patterns in the relationship between irAE severity, cancer type, and treatment regimen with respect to both irAE-free survival and overall survival. In patients with severe irAEs, both time to irAE event and overall survival were significantly affected, highlighting the dual impact of irAE severity on clinical outcomes. In contrast, among multi-site cancer patients, the treatment regimen significantly influenced irAE-free survival but did not translate into differences in overall survival. Conversely, in kidney cancer patients, the treatment regimen significantly impacted overall survival but had a limited effect on irAE-free survival. These findings suggested that the relationship between treatment regimen and survival outcomes is highly context-dependent, varying by cancer type and endpoint. Future studies should focus on therapeutic strategies tailored to these specific contexts to optimize the safety and efficacy of ICI therapies.

While our study provided valuable insights, several limitations must be acknowledged. First the observed associations may be influenced by unmeasured or residual confounding factors, which we could not fully control despite adjusting for several potential confounders such as age, sex, and comorbidities. However, genetic predispositions and lifestyle factors, including diet, physical activity, and environmental exposures, play a critical role in modulating the immune system’s response to ICIs but were not captured in our analysis. Future studies incorporating genetic profiling and detailed lifestyle data are necessary to comprehensively assess these influences. Additionally, the relatively small sample size in certain subgroups, such as patients with less common cancer types and those receiving specific ICI regimens, may have limited the statistical power of our analyses. This could result in non-significant findings in some areas, such as the irAE-free survival analysis for patients with kidney and multi-site cancers, despite potentially meaningful clinical trends. Larger studies would allow for more robust subgroup analyses and greater confidence in the observed associations. Another important limitation is our inability to differentiate between varying severities of irAEs. Some irAEs are mild and self-limiting, while others are severe, requiring intervention or leading to life-threatening outcomes. Our study treats all irAEs as a homogeneous outcome, which overlooks the nuanced impact that irAE severity may have on patient prognosis and treatment continuation. Future research should focus on stratifying irAEs by severity to provide a more detailed understanding of how different grades of irAEs influence clinical outcomes, therapeutic decisions, and quality of life for patients undergoing ICI therapy.

## CONCLUSIONS

This study provides critical insights into the factors influencing the development of irAEs in patients undergoing ICI therapy. By identifying the key demographic, clinical, and treatment-related predictors of irAEs, our findings offer valuable guidance for clinicians in tailoring ICI treatments to individual patients, particularly in populations at higher risk for adverse events. Furthermore, this study emphasizes the need for early identification and management of irAEs to improve patient outcomes. As ICIs continue to play an increasingly central role in cancer treatment, these insights will contribute to optimizing therapy and mitigating risks across diverse patient populations. By expanding on these findings and addressing the limitations outlined, future research can further advance our understanding of irAEs and improve the safety and efficacy of ICI therapies in clinical practice.

## Data Availability

All data produced in the present study are available upon reasonable request to the authors.

## Conflict of Interest Disclosures

The authors have no conflict of interest to disclose.

### Abbreviations

ICI: immune checkpoint inhibitors ir
AE: immune-related adverse events

