## Supplementary for "A real-world cohort study of immune-related adverse events in patients receiving immune checkpoint inhibitors"

**Supplementary Figure 1. Algorithm for phenotyping irAE**

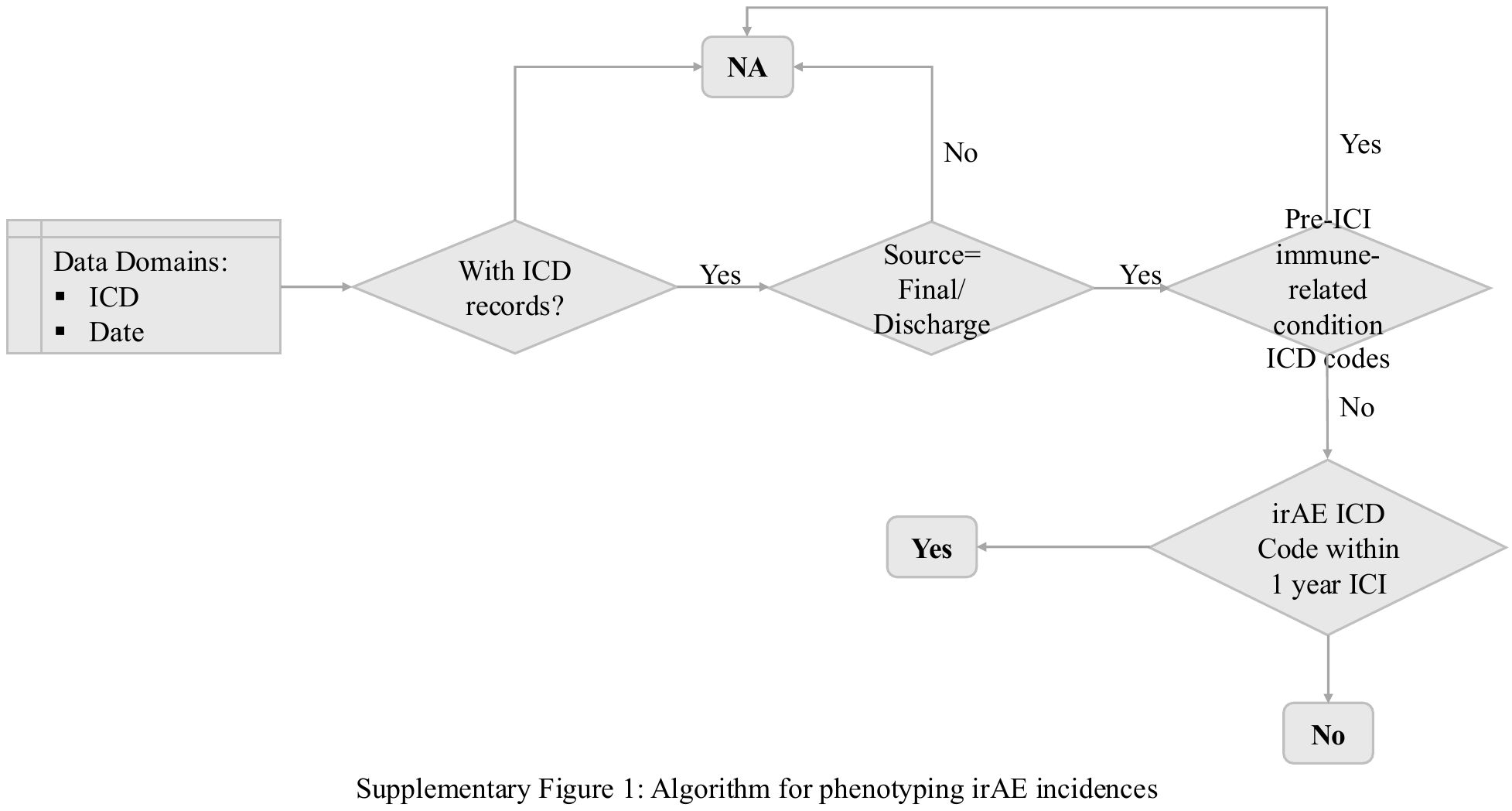

**Supplementary Figure 2. irAE risk evaluation with demographic features**

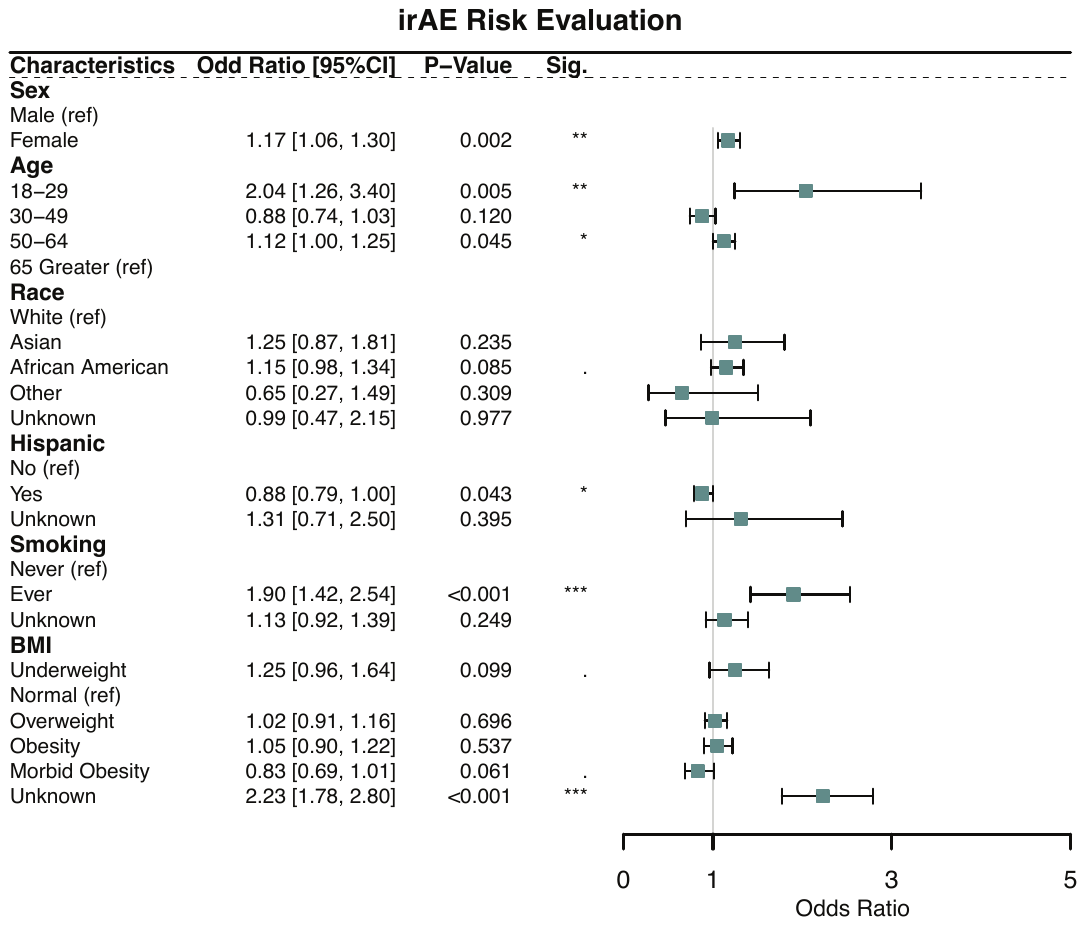

**Supplementary Figure 3. irAE risk evaluation with comorbidity**

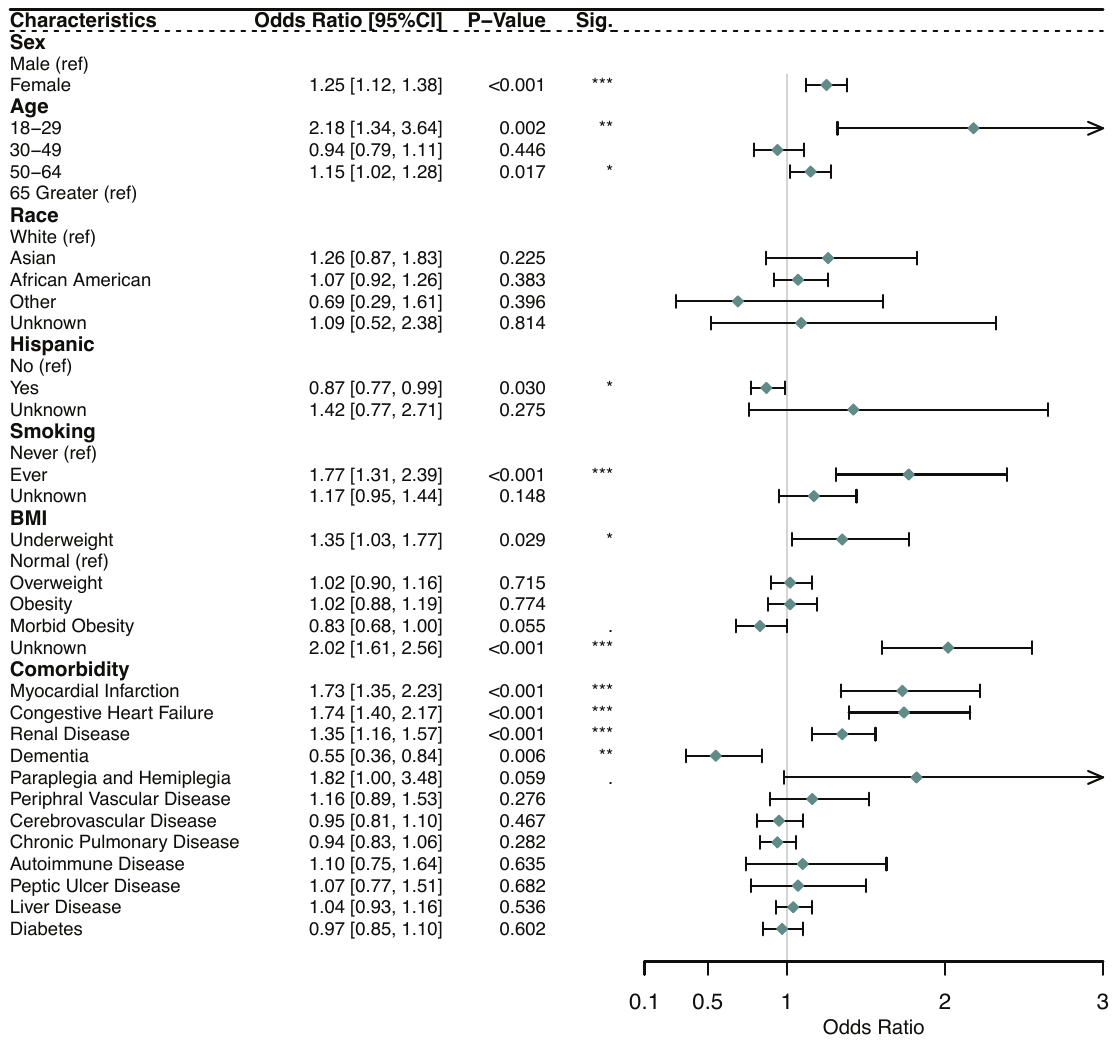

**Supplementary Figure 4. irAE risk evaluation with ICI treatment**

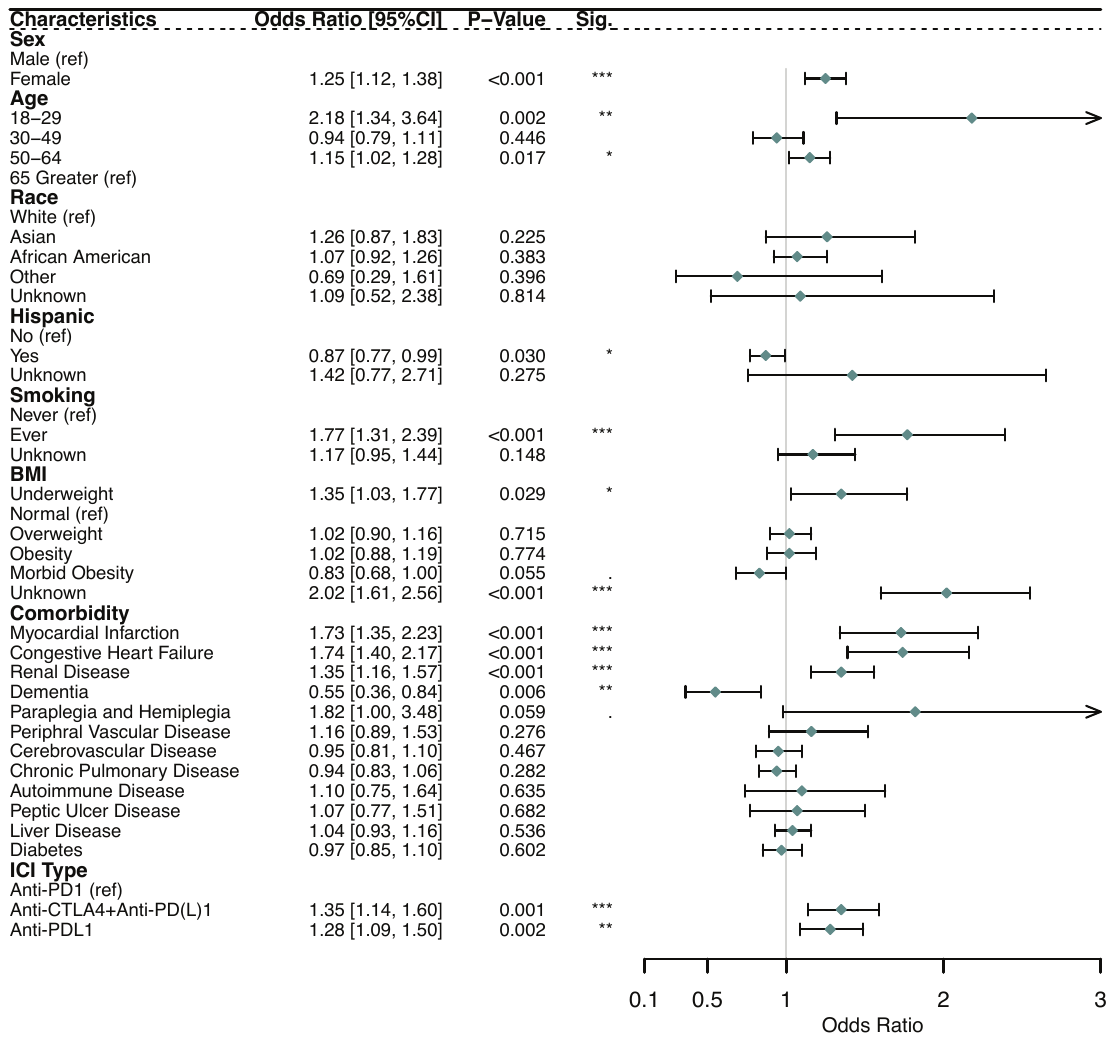

**Supplementary Figure 5. irAE risk evaluation excluding gender-specific irAE**

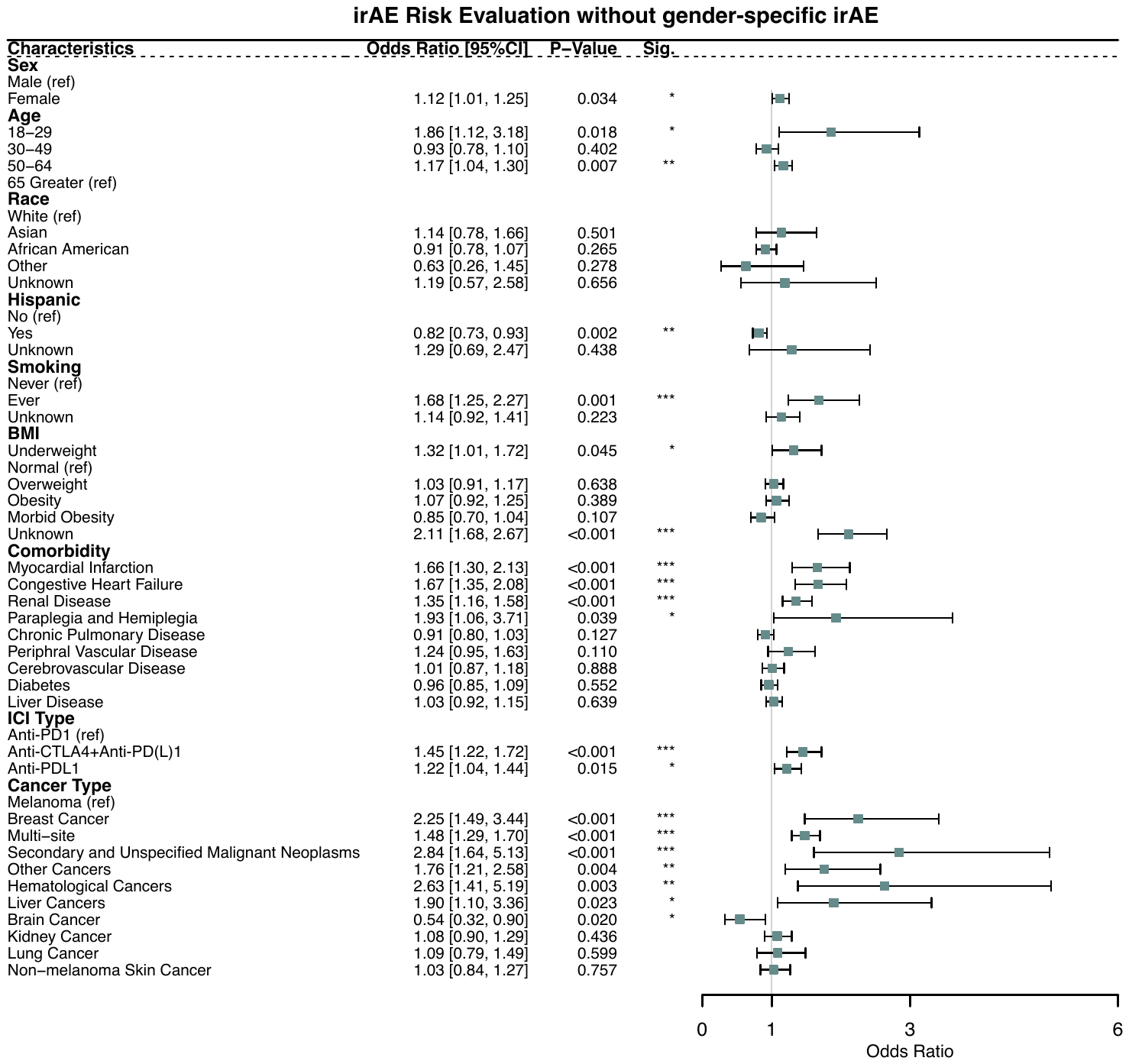

**Supplementary Figure 6. Kaplan-Meier Analysis**

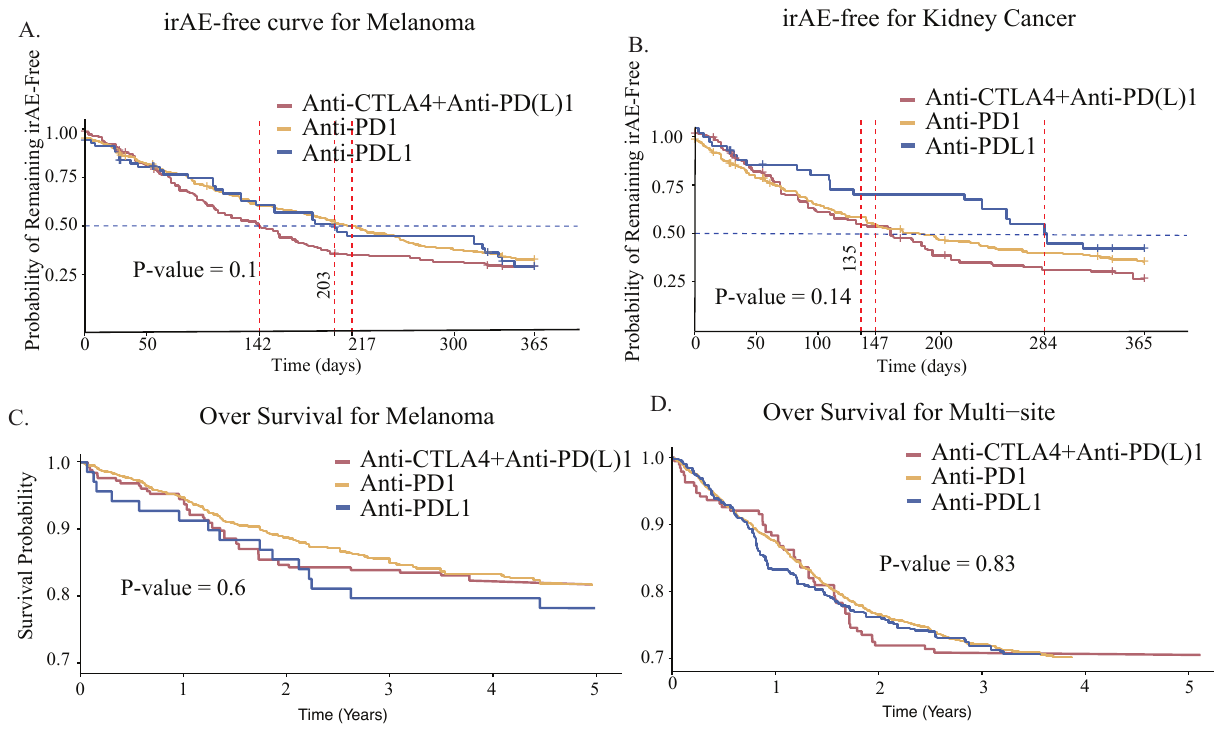

**Supplementary Figure 7. Cumulative incidence curve of irAE**

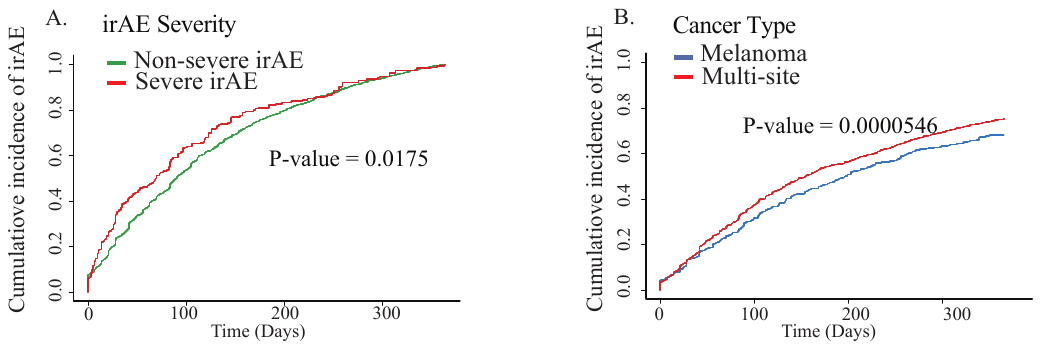

**Supplementary Table 1. Cohort characteristics of patients receiving ICI with expanded cancer type**

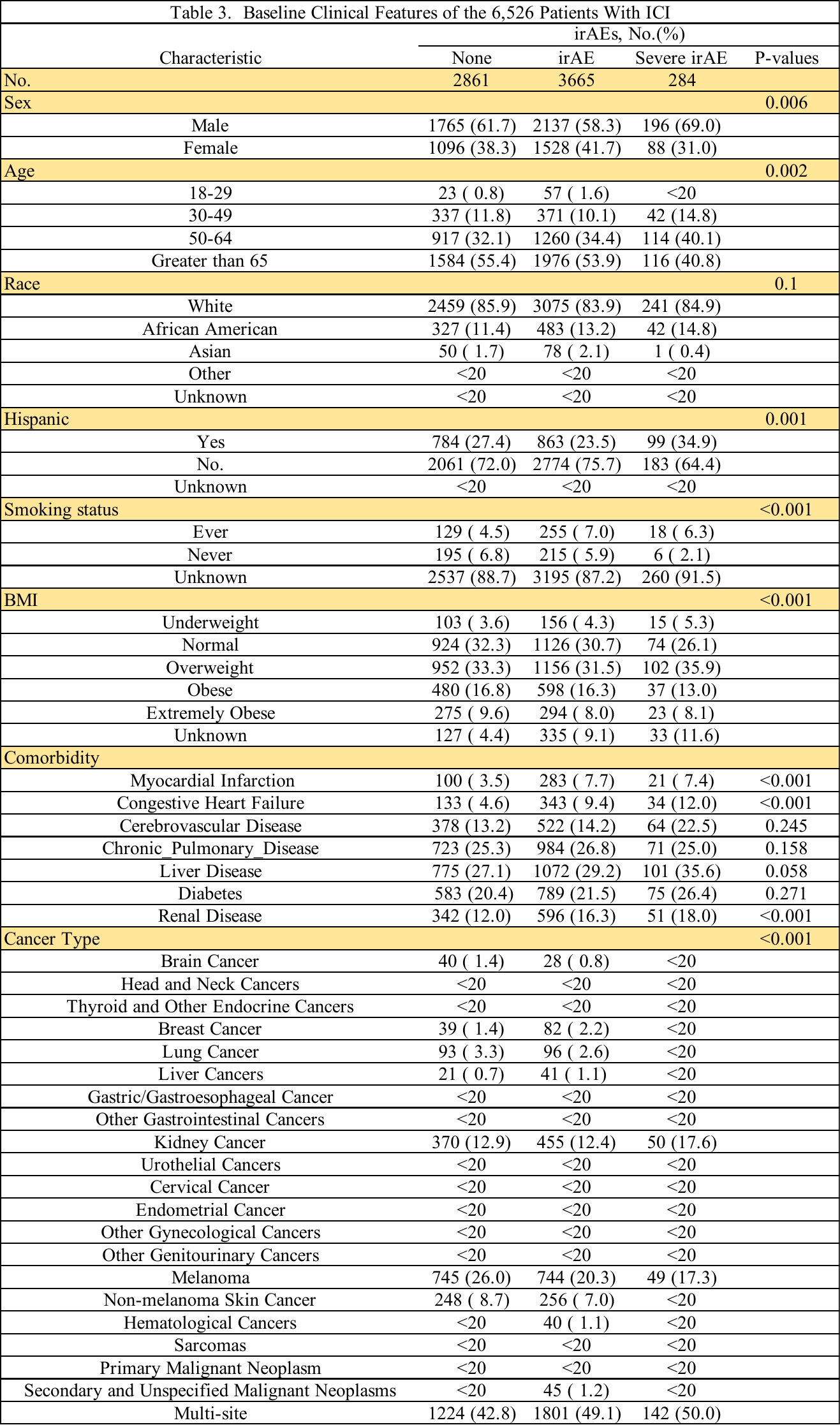

**Supplementary Table 2. Cohort characteristics of irAE patients**

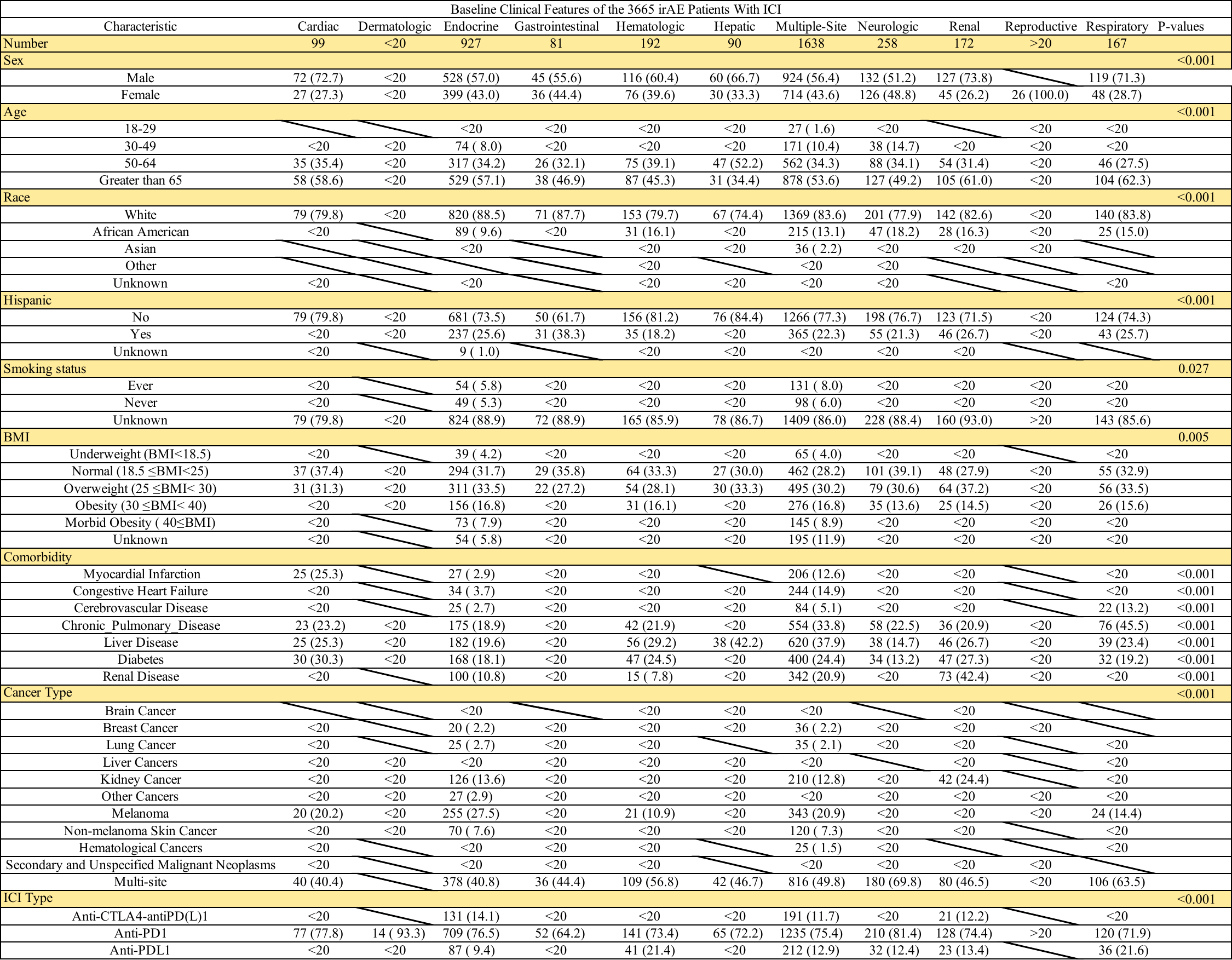
